# Long working and commuting times as risk factors for depressive symptoms. Cross-sectional and longitudinal analyses

**DOI:** 10.1101/2022.12.22.22283831

**Authors:** Nico Dragano, Hermann Burr, Maren Formazin, Anika Schulz, Uwe Rose

## Abstract

**Background:** Regular long working and commuting hours are thought to have negative consequences for mental health. However, the study results are not clear and vary by country. The present analysis examines associations between working or commuting hours and depressive symptoms for Germany.

**Method:** The S-MGA study (German Study on Mental Health at Work) is a longitudinal cohort of a random sample of employees subject to social insurance contributions. We analysed data from 3 413 participants of the baseline survey (cross-sectional analysis) and from 2 019 people who participated at baseline and at a follow-up survey five years later (longitudinal analysis). Weekly working and commuting hours as well as covariates (age, gender, occupational position, psychosocial working conditions) were collected at baseline. Depressive symptoms were recorded with the *Patient Health Questionnaire* at both waves. Multivariate logistic regression models were used to control for covariates.

**Results:** At baseline survey, 7 % of the employees had overlong working hours of ≥ 55 hours per week, and another 8 % worked > 48-54 hours. Long working hours were cross-sectionally associated with moderately elevated depressive symptoms compared to normal working hours (35-< 40 h/week). When new depressive symptoms after five years were considered, the correlation was significant for > 55 weekly working hours (odds ratio [OR] 2.14; 95 % confidence interval [CI] 1.11;4.12), but not for > 48-54 h (OR 1.26, CI 0.65;2.43). Employees who commuted ten hours or more per week had more depressive symptoms cross-sectionally (OR 1.83; CI 1.13;2.94) compared to the reference group who commuted < 2.5 hours. This correlation was not observed longitudinally.

**Conclusions:** The results suggest that excessive working and commuting time is associated with depressive symptoms in employees, although the effects of commuting time were only found cross-sectionally. The results underline the importance of adhering to working time regulations and avoiding excessive working hours. Further research is needed on the role of commuting.

## Background

The identification of work-related risk factors for mental illness is important in view of the high number of mental illnesses among employees (1) (2). One factor discussed in this context is long working hours. They are defined mostly as a significant deviation from normal working hours, which are usually set at 35 to < 41 hours per week. The International Labour Organisation (ILO) refers to long working hours as starting from 49 hours per week (3). International research has also established a limit for excessive working hours of 55 hours or more (4, 5).

Working weeks of 49 up to 60 hours are permitted in Germany for a short period of time if they are subsequently compensated for by shorter working weeks (3). Even though the average working time in this country has been decreasing for years, the proportion of employees who regularly work more than 48 hours is considerable (6): for example, 9.7 % of employees reported regular weekly working hours of more than 48 hours in 2019 (7). However, there is a variation depending on occupation and position. The share among managers, for example, is 30.3 %, and among the self-employed with employees it is as high as 50.7 % (7). The health sector should also be highlighted, where long working hours are more common than in other sectors (8).

Long working hours can impair the necessary recuperation after mental and physical stress during work (10). Long working hours may also have negative effects on health-related behaviour, as they limit the opportunities for health-promoting activities during leisure time, such as sports (11). People’s “work-life balance” is also affected: long working hours are cross-sectionally associated with conflicts between work and private life and psychosomatic complaints (12-14). The less time available for maintaining social contacts and relationships, the higher the probability of conflicts and negative feelings (15). In addition, occupations with long working hours are often those with high overall quantitative demands, which means that long working hours are also completed under high psychological stress (10).

For these reasons, long and excessive working hours are considered a potential risk factor for the development of depressive symptoms and major depression (5). The evidence for an association is based on a limited number of long-term epidemiological studies, and their results are inconsistent, as two recent reviews and meta-analyses on the topic show. Virtanen and colleagues report a pooled relative risk of 1.14 (95 % confidence interval 1.03;1.25) for the onset of depressive symptoms when employees work more than 55 hours per week (reference: 35 – 40 h) (4). Based on these findings, the World Health Organization decided to include excessive working hours in the list of factors used to calculate the global burden of disease (16) and initiated a corresponding meta-analysis (5). In this analysis, 22 longitudinal studies were summarised, and the result was that excessive working hours (55 or more hours per week) were not significantly associated with the incidence of depression (5). However, both reviews assessed the quality of the studies included as low (4, 5). In addition, the studies were from only a few countries, with a focus on the USA and Scandinavia. Both reviews also noted that the results of the individual studies were very different, so that combining them into a common estimate should be considered critical. It is, therefore, conceivable that there are country-specific differences in working conditions or occupational health and safety legislation that moderate the possible effects of working hours on health (17). Only results from two longitudinal studies based on the Socio-Economic Panel (SOEP) and the Pairfam Panel have been reported for Germany so far, which were evaluated in Virtanen’s meta-analysis (4). The analyses based on the SOEP show a moderate increase in risk, however, no association between working hours and depressive symptoms could be identified in the small study population of the Pairfam study.

Another aspect that has hardly been taken into consideration so far is that pure working time does not fully reflect the time spent on gainful employment, as the commute is not taken into account. Commuting is a widespread phenomenon; in 2017, 69 % of all employees commuted at least 2.5 hours per week (18). Despite the trend towards more work from home (e.g. teleworking, working from home), which saw a surge in the COVID-19 pandemic, occupational commuting will also exist in the future for at least some of the employees due to the nature of their work, for example, in trades, industry and personal services. The literature reports contradictory results regarding commuting time and indicators of mental health. A cross-sectional study with employees in an industrial company in Germany found negative associations of commuting time with mental health (20), as did a longitudinal study in the UK (21). By contrast, a longitudinal study from Australia showed moderately increased long-term effects (19).

Another important issue when investigating effects of work and commuting time is the length of follow-up time in longitudinal studies of depressive symptoms (22). Current findings from research on psychosocial workload show that the associations between stress and mental health vary depending on the time perspective (23).

Evidence on the relationship between excessive working and commuting times and depressive symptoms is limited, especially for Germany. Therefore, this study will investigate this topic using data from a cohort study of employees subject to social insurance contributions in Germany, both cross-sectionally and longitudinally.

## Methods

### Study Sample

The analysis is based on data from the “Study on Mental Health at Work” (*Studie Mentale Gesundheit bei der Arbeit*: S-MGA), a longitudinal study on the mental health of employees in Germany conducted by the Federal Institute for Occupational Safety and Health (BAuA) (24). The sample was recruited based on a random selection of employees subject to social insurance contributions (31 – 60 years). A total of 4 511 people participated in the baseline survey conducted in 2011/2012 (AAPOR participation rate = 36 %) (25). All respondents were informed about the study objectives and declared their willingness to participate in writing (24). The analytical sample for this analyses excluded a number of non-eligible subjects. First, respondents who had given up their employment subject to social security contributions in the period between the sampling and the survey were excluded (n = 310; Figure 1). In addition, we excluded 269 people who were only employed on a marginal scale of less than ten hours per week and 519 respondents who had missing data for at least one of the study variables. Data from 3 413 people were, thus, available for the cross-sectional analyses of the baseline survey. Of these, 2 119 participated in the follow-up survey five years later (AAPOR participation rate at follow-up = 69 %). One hundred people had missing scores on the scale measuring depressive symptoms and could not be included in these analyses. This left 2 019 participants for the longitudinal analysis.

**Figure 1:**
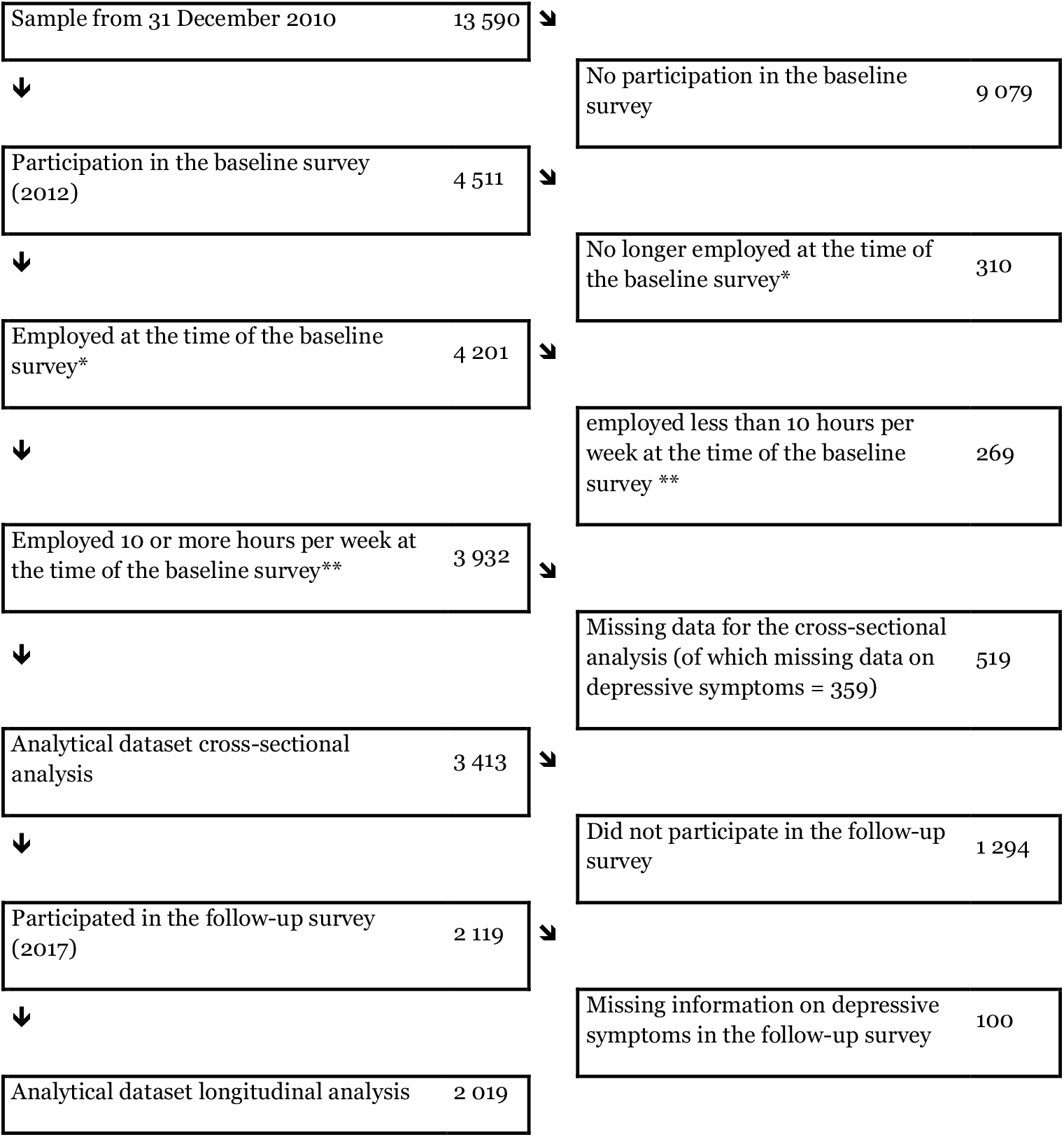
Flow chart – from the sample drawn in 2010 to the cross-sectional (2012) and longitudinal (2012–2017) analysis sample of the S-MGA. ^*^ The average time between the sampling date and the baseline survey was 13 months (range: 9 – 17). A total of 310 people who were employed at the time of sampling had stopped working by the time of the baseline survey and were excluded. ^**^ Weekly working hours in main and secondary employment.

### Data Collection

Data collection was carried out at both survey waves with computer-assisted personal interviews (CAPI), which were conducted by trained interviewers in the participants’ homes using a standardised questionnaire. Following the interview, the subjects completed a questionnaire on depressive symptoms in the absence of the interviewers in order to minimise survey effects, for example, socially desirable answers during interviews (26). The study received a positive ethics approval from the ethics committee of the BAuA (Votum 006_2016_Müller).

### Depressive Symptoms

The outcome variable of this analysis was measured with the German-language version of the “Patient Health Questionnaire” (PHQ-9). The latter is a screening instrument designed for use in clinical settings and assesses the presence of the main symptoms of major depression according to DSM-IV criteria (27, 28). Some examples of symptoms are “little interest or pleasure in doing things” or “feeling down, depressed or hopeless”. The frequency with which the symptoms occurred in the 14 days before the interview [(0) not at all; (1) on single days; (2) on more than half of the days; (3) almost every day] was recorded. The total score was determined as the sum of all item scores and can take values between 0 and 27. The presence of depressive symptoms was determined with a cut-off value of ≥ 10 (27, 29).

### Excessive working hours and commuting

The number of hours worked was surveyed for both the main job and any secondary jobs as average weekly working hours in an open response format. The total number of hours worked per week for all activities was divided into the categories commonly used in international research: ‘10 to less than 35 hours’, ‘35 to less than 41 hours’ (reference category), ‘41 to less than 49 hours’, ‘49 to less than 55 hours’ and ‘55 and more hours’ (5, 30). Time spent commuting to and from work each day was measured in minutes and converted to the total weekly time analogous to working time. In contrast to working time, there is no internationally established classification for commuting times. Therefore, a scale developed by the BAuA was used (“less than 2.5 hours”, “2.5 to less than 5 hours”, “5 to less than 10 hours” and “10 and more hours”) (18).

### Control and stratifying variables

Age and gender were included as control variables. Age was used with three categories: 31 to 40, 41 to 55, and 56 to 60 years, in order to make non-linear correlations between depressive symptoms and age discernible (31, 32). Occupational position was recorded based on the current occupation coded according to the “International Standard Classification of Occupations (ISCO-08)” with four categories according to the classification of the level of education “International Standard Classification of Education (ISCED)”: unskilled workers (ISCO major group 9), professionals (4-8), senior professionals (3) and academic and managerial occupations (1,2) (33, 34). Managerial occupations are not considered in ISCED, which focuses on educational attainment. For the purpose of this paper, managers were grouped together with academic occupations in one ISCED group (35).

A possible confounder in the relationship between working hours and depressive symptoms is the extent of psychological stress during working hours. Two scales from the Copenhagen Psychosocial Questionnaire (COPSOQ) were used to control for this (36). They represent the two central dimensions of the demand-control model (37). “Quantitative work demands” are measured by the mean value of the answers to five items on the amount and pace of work, where values between 0 and 4 are possible and high values indicate high stress. “Control over one’s own work” was surveyed as the mean value of the answers to four items on influence at work. This indicator can also take on values between 0 and 4, with high values indicating a high level of control over work in a positive sense.

Potential changes of employer were identified in the follow-up survey: respondents could indicate whether they had changed employers since the baseline survey (coded as change no/yes).

### Statistical Methods

Multiple logistic regressions were calculated separately for both exposures in order to statistically estimate the influence of excessive working and commuting times (independent variables) on the occurrence of depressive symptoms (dependent variable). Both cross-sectional correlations and time-lagged effects were considered (22, 23). In addition to models without adjustment (model 1), adjusted models were calculated. In the cross-sectional analyses we included the control variables age and gender (model 2), as well as occupational position, quantitative demands and control (model 3). For the longitudinal analyses we first adjusted for depressive symptoms at baseline (model 2). Then age and gender (model 3), and occupational position, quantitative demands and control (model 4) were added.

In addition, we investigated whether the joint incidence of long working and commuting times was particularly associated with depressive symptoms. For this purpose, the variables working and commuting times were dichotomised (for working time: less than 49 hours vs. 49 hours and more; for commuting time: up to 5 hours vs. 5 hours and more) and four groups were formed. The presence of an over-additive interaction was tested by calculating the “relative excess risk due to interaction” (RERI) (38). The results of the corresponding logistic regressions are presented as odds ratios (ORs) with 95 % confidence intervals (CI) and, in the case of the RERI calculation, as relative risks (RRs) (38).

Three sensitivity analyses were conducted to validate the results. In the first sensitivity analysis, the cross-sectional and longitudinal effects were determined separately by gender in order to check whether results are comparable for women and men. Longitudinal analyses were then calculated separately for people who worked for the same employer during the five-year study period (n = 1 488) and those who no longer worked for the same employer (n = 531). The first group can be assumed to have a rather constant exposure during the study period compared to the latter group. In the third sensitivity analysis, the cross-sectional and longitudinal effects were determined separately by occupational position (unskilled workers and skilled workers, n = 940; senior skilled workers, academic professions and managers, n = 1 079) in order to determine their influence.

All analyses were carried out with the statistical programme SPSS IBM version 27.

## Results

### Description of the Population

Men and women were equally represented in the sample of 3 413 employees in the baseline survey. The majority of them were older than 40 years and approximately 50 % worked in higher professional positions (Table 1). Mean weekly working time in the sample was 38.9 hours and employees commuted an average of 3.9 hours a week. Taken together, 42 % of the respondents worked 41 hours or more per week. While 8 % worked 49 to less than 55 hours per week, a working week of 55 hours or more was reported by 7 % of respondents. Long commuting times of 10 or more hours per week were also reported by 7 % of the employees. The correlation between working and commuting times was low with r = 0.104 (table not shown). Table 1 also shows the mean working and commuting times for individual subgroups. Thus, on average, men worked longer hours than women (men = 44.4 hours, women = 33.4 hours) and men’s commuting times were also longer (men = 4.2 hours, women = 3.6 hours). Regarding occupational position, employees in higher occupational positions tended to be more affected by long working and commuting times.

**Table 1:** Characteristics of the baseline survey sample (N = 3 413)

|  |  | N | % | M (SD) | Average working time<br>M (SD) | Average<br>commuting time<br>M (SD) |
| --- | --- | --- | --- | --- | --- | --- |
| CASES |  | 3 413 | 100 | - | 38.9 (11.8) | 3.9 (3.2) |
| GENDER | Male | 1 697 | 50 | - | 44.4 (8.5) | 4.2 (3.5) |
|  | Female | 1 716 | 50 | - | 33.4 (12.0) | 3.6 (2.9) |
| AGE | 31 – 40 | 840 | 25 | - | 40.0 (12.1) | 4.0 (3.4) |
|  | 41 – 55 | 2 060 | 42 | - | 38.3 (11.7) | 3.9 (3.1) |
|  | 56 – 60 | 513 | 34 | - | 38.7 (11.5) | 4.0 (3.3) |
| OCCUPATIONAL POSITION | Unskilled workers | 206 | 6 | - | 31.4 (13.3) | 3.3 (2.9) |
|  | Professionals | 1 521 | 45 | - | 38.3 (11.9) | 3.6 (3.1) |
|  | Senior professionals | 915 | 27 | - | 37.6 (10.2) | 4.3 (3.4) |
|  | Academics and managerial occupations | 771 | 23 | - | 43.5 (11.8) | 4.4 (3.4) |
| QUANTITATIVE DEMANDS* |  | 3 413 | - | 2.3 (0.8) | - | - |
| CONTROL* |  | 3 413 | - | 2.2 (0.7) | - | - |
| WEEKLY WORKING HOURS |  | 3 413 | - | 38.9 (11.8) | - | - |
|  | 10 to < 35 hours | 836 | 24 | - | 22.6 (7.0) | 3.3 (2.7) |
|  | 35 to < 41 hours | 1 113 | 33 | - | 38.7 (1.7) | 4.1 (3.2) |
|  | 41 to < 49 hours | 925 | 27 | - | 44.0 (2.0) | 4.3 (3.5) |
|  | 49 to < 55 hours | 289 | 8 | - | 50.4 (1.0) | 4.0 (3.6) |
|  | ≥ 55 hours | 250 | 7 | - | 61.6 (7.3) | 4.0 (3.6) |
| WEEKLY COMMUTING TIME |  | 3 413 | - | 3.9 (3.2) | - | - |
|  | < 2.5 hours | 1 071 | 31 | - | 37.4 (13.3) | 1.2 (0.5) |
|  | 2.5 to < 5 hours | 1 268 | 37 | - | 38.8 (11.3) | 3.2 (0.6) |
|  | 5 to <10 hours | 826 | 24 | - | 40.0 (10.3) | 6.1 (1.2) |
|  | ≥ 10 hours | 248 | 7 | - | 41.8 (10.9) | 12.3 (4.1) |
| DEPRESSIVE SYMPTOMS<br>(PHQ-9) | Average† | 3 413 | - | 4.3 (3.5) | - | - |
|  | No (PHQ <10) | 3 144 | 92 |  | 38.8 (11.8) | 3.9 |
|  | Yes (PHQ ≥10) | 269 | 8 |  | 39.5 (11.8) | 4.3 |

All percentages rounded; therefore, they do not always add up to 100. M = Mean, SD = Standard Deviation
\* The scale ranges from 0 to 4. † The scale ranges from 0 to 27.

Both working and commuting times in the cross-sectional analyses were – with one exception – not significantly associated with high depressive symptoms (Table 2). A significantly increased OR was only observed in the group of employees with commuting times of 10 or more hours per week (model 3: OR = 1.83, 95 % CI = 1.13;2.94). The OR was increased for excessive working hours of 55 and more hours, but the CI included 1 (model 3: OR = 1.45, 95 % CI = 0.83;2.51).

**Table 2:**
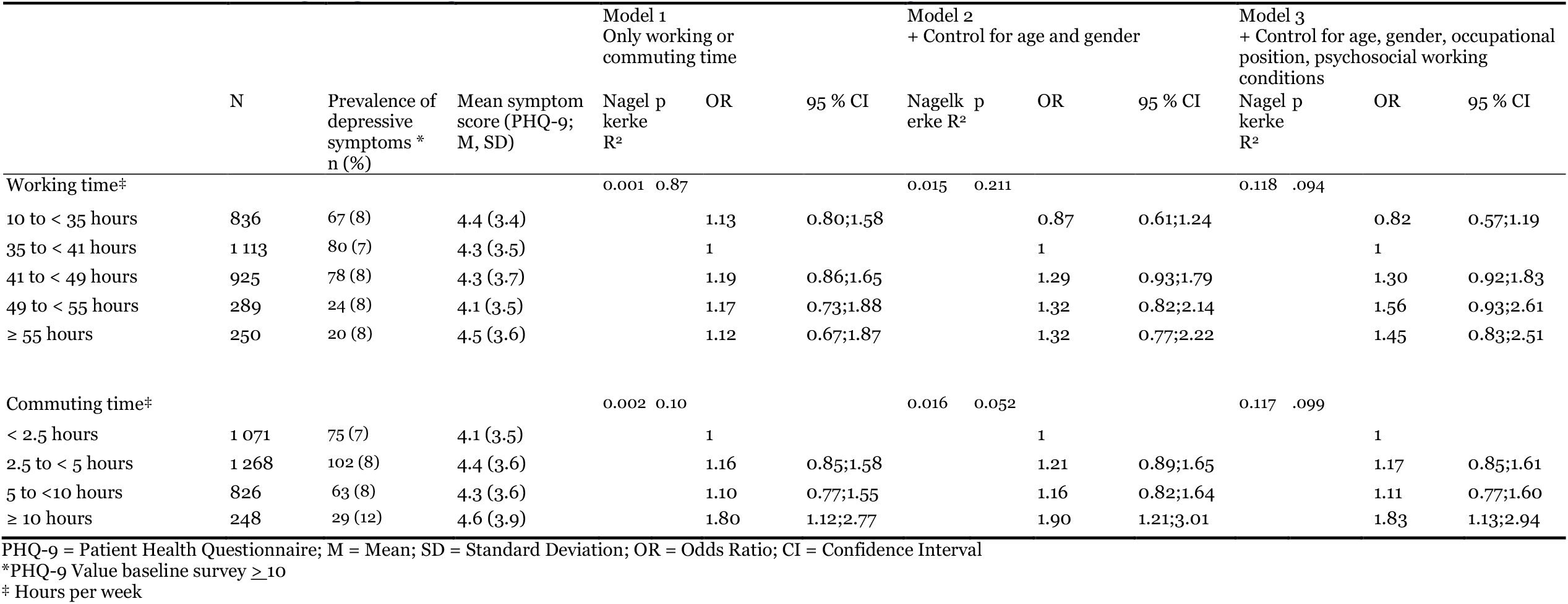
Associations (cross-sectional) between work and commuting time and depressive symptoms in 3 413 employees of the S-MGA baseline survey (logistic regression; odds ratios (OR) and 95 % confidence intervals (CI)).

Table 3 shows the results of the longitudinal analysis. Employees with the longest working hours (55 ≥ hours) had a 2.14-fold increased OR (95 % CI = 1.11;4.12) of reporting depressive symptoms after five years compared to employees with working hours of 35 to < 41 hours in the fully adjusted model 4. The ORs were increased although not statistically significant for weekly working hours of 41 to < 49 and 49 to < 55 hours (OR = 1.34, 95 % CI = 0.88;2.03 and OR = 1.26, 95 % CI = 0.65;2.43, respectively). No effect was observed for commuting times: Employees with particularly long commuting times even tended to have a lower risk of depressive symptoms than those with low commuting times of less than 2.5 hours per week, but the association was weak in all cases.

**Table 3:** Associations (longitudinal) between working and commuting times at baseline and depressive symptoms at follow-up in 2 019 S-MGA employees (logistic regression; odds ratios (OR) and 95 % confidence intervals (CI)).

|  | N | Depressive symptoms at the follow-up survey | Model 1<br>Only working or commuting time |  |  |  | Model 2<br>+ Control for depressive symptoms at baseline |  |  |  | Model 3<br>+ Control for depressive symptoms at baseline, age and gender |  |  |  | Model 4<br>+ Control for depressive symptoms at baseline, age, gender, occupational position, psychosocial working conditions |  |  |  |
| --- | --- | --- | --- | --- | --- | --- | --- | --- | --- | --- | --- | --- | --- | --- | --- | --- | --- | --- |
|  |  | Prevalence*<br>n (%) | Nagelkerke<br>R<sup>2</sup> | p | OR | 95 % CI | Nagelkerke<br>R<sup>2</sup> | p | OR | 95 % CI | Nagelkerke<br>R<sup>2</sup> | p | OR | 95 % CI | Nagelkerke<br>R<sup>2</sup> | p | OR | 95 % CI |
| Working time‡ |  |  | 0.001 | 0.811 |  |  | 0.147 | 0.745 |  |  | 0.171 | 0.155 |  |  | 0.182 | 0.062 |  |  |
| 10 to < 35 hours | 496 | 48 (10) |  |  | 1.04 | 0.70;1.54 |  |  | 1.08 | 0.71;1.64 |  |  | 0.83 | 0.53;1.28 |  |  | 0.80 | 0.52;1.25 |
| 35 to < 41 hours | 673 | 63 (9) |  |  | 1 |  |  |  | 1 |  |  |  | 1 |  |  |  | 1 |  |
| 41 to < 49 hours | 538 | 57 (11) |  |  | 1.15 | 0.79;1.67 |  |  | 1.13 | 0.76;1.69 |  |  | 1.28 | 0.85;1.93 |  |  | 1.34 | 0.88;2.03 |
| 49 to < 55 hours | 172 | 15 (9) |  |  | 0.92 | 0.51;1.67 |  |  | 0.96 | 0.52;1.80 |  |  | 1.13 | 0.60;2.13 |  |  | 1.26 | 0.65;2.43 |
| ≥ 55 hours | 140 | 17 (12) |  |  | 1.34 | 0.76;2.37 |  |  | 1.49 | 0.82;2.73 |  |  | 1.82 | 0.97;3.38 |  |  | 2.14 | 1.11;4.12 |
| Commuting time‡ |  |  | 0.002 | 0.607 |  |  | 0.148 | 0.507 |  |  | 0.166 | 0.755 |  |  | 0.175 | 0.614 |  |  |
| < 2.5 hours | 613 | 68 (11) |  |  | 1 |  |  |  |  |  |  |  | 1 |  |  |  | 1 |  |
| 2.5 to < 5 hours | 758 | 74 (10) |  |  | 0.87 | 0.61;1.23 |  |  | 0.83 | 0.58;1.21 |  |  | 0.87 | 0.60;1.27 |  |  | 0.83 | 0.57;1.21 |
| 5 to < 10 hours | 498 | 46 (9) |  |  | 0.82 | 0.55;1.21 |  |  | 0.80 | 0.53;1.22 |  |  | 0.86 | 0.56;1.31 |  |  | 0.82 | 0.53;1.25 |
| ≥ 10 hours | 150 | 12 (8) |  |  | 0.70 | 0.37;1.32 |  |  | 0.63 | 0.32;1.25 |  |  | 0.72 | 0.36;1.43 |  |  | 0.68 | 0.34;1.36 |

PHQ-9 = Patient Health Questionnaire; M = Mean; SD = Standard Deviation; OR = Odds Ratio; CI = Confidence Interval
\*PHQ-9 Value baseline survey ≥10
‡ Hours per week

The question of whether it is particularly disadvantageous if employees have both long working and commuting times was answered using an interaction analysis (Table 4). This tended to be the case for both the cross-sectional and longitudinal analysis. However, the RERI, which indicates the presence of an over-additive interaction, was not significant in either case.

**Table 4:**
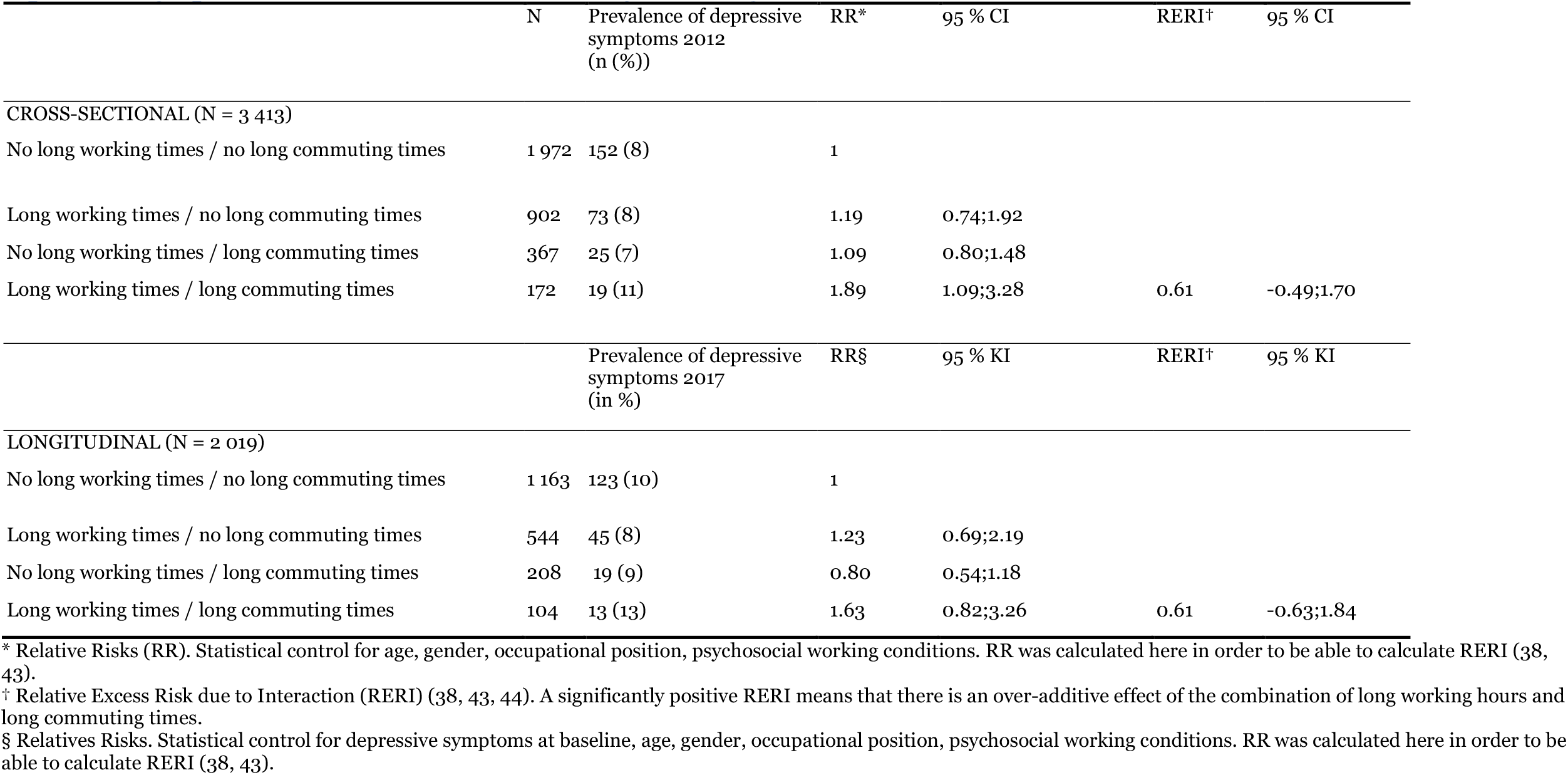
Interaction of long working time (≥ 49 hrs. week) and long commuting times (≥ 5 hrs. week) for the occurrence of depressive symptoms in the cross-section and longitudinally.

|  | N | Prevalence of depressive symptoms 2012<br>(n (%)) | RR* | 95 % CI | RERI† | 95 % CI |
| --- | --- | --- | --- | --- | --- | --- |
| CROSS-SECTIONAL (N = 3 413) |  |  |  |  |  |  |
| No long working times / no long commuting times | 1 972 | 152 (8) | 1 |  |  |  |
| Long working times / no long commuting times | 902 | 73 (8) | 1.19 | 0.74;1.92 |  |  |
| No long working times / long commuting times | 367 | 25 (7) | 1.09 | 0.80;1.48 |  |  |
| Long working times / long commuting times | 172 | 19 (11) | 1.89 | 1.09;3.28 | 0.61 | -0.49;1.70 |
|  |  | Prevalence of depressive symptoms 2017<br>(in %) | RR§ | 95 % KI | RERI† | 95 % KI |
| LONGITUDINAL (N = 2 019) |  |  |  |  |  |  |
| No long working times / no long commuting times | 1 163 | 123 (10) | 1 |  |  |  |
| Long working times / no long commuting times | 544 | 45 (8) | 1.23 | 0.69;2.19 |  |  |
| No long working times / long commuting times | 208 | 19 (9) | 0.80 | 0.54;1.18 |  |  |
| Long working times / long commuting times | 104 | 13 (13) | 1.63 | 0.82;3.26 | 0.61 | -0.63;1.84 |

\* Relative Risks (RR). Statistical control for age, gender, occupational position, psychosocial working conditions. RR was calculated here in order to be able to calculate RERI (38, 43).
† Relative Excess Risk due to Interaction (RERI) (38, 43, 44). A significantly positive RERI means that there is an over-additive effect of the combination of long working hours and long commuting times.
§ Relatives Risks. Statistical control for depressive symptoms at baseline, age, gender, occupational position, psychosocial working conditions. RR was calculated here in order to be able to calculate RERI (38, 43).

### Sensitivity analyses

The first sensitivity analysis examined whether the effects of working and commuting time on depressive symptoms vary by gender (Online Appendix Tables A1 and A2). No gender differences were found, with two exceptions. The risk of depressive symptoms was increased for men and reduced for women when working less than 35 hours. The risk was higher for men than for women for commuting times of 10 hours or more. The second sensitivity analysis examined whether the effects of working and commuting time on depressive symptoms in the longitudinal section depended on whether the employees changed their employer or not (Online Appendix Table A3). In general, no significant differences were found between the subgroups, but with two exceptions: for working hours of less than 35 hours and commuting times of 2.5 to < 5 hours. A further stratified analysis by occupational position showed no differences between the subgroups (Online Appendix Tables A4 & A5).

## Discussion

The results suggest that excessive working hours and commuting times were differentially associated with depressive symptoms among women and men in Germany who are subject to social insurance contributions. In the case of excessively long working hours (55 hours and more), pronounced effects were found in the longitudinal analyses, whereas in the case of long commuting times (10 hours and more), associations were present only in the cross-sectional analyses. A workweek of 55 hours or more revealed more than twice the longitudinal risk of depressive symptoms; commuting times of 10 hours or more nearly doubled the cross-sectional prevalence. When working hours ranged from 41 to less than 55 hours, we found a slight increase in depressive symptoms in both the cross-sectional data and the longitudinal data. However, there were indications that there might be an additional effect on depressive symptoms among those employees who have both long working hours and long commuting times.

The result that excessive working hours were longitudinally associated with depressive symptoms among women and men in Germany corresponds with the results of an analysis of the SOEP study, which is also based on a representative sample in Germany (4). However, it contrasts with an analysis of the German Pairfam study, which found no significant effects based on a smaller sample of younger workers in Germany (4). As was mentioned in the introduction, there are also contradictory results internationally regarding the association between long working hours and depressive symptoms. The two most recent reviews and metaanalyses have shown that the results of the individual studies are indeed inconsistent (4, 5) and that more studies from individual countries are needed to draw valid conclusions. The fact that clear longitudinal correlations were discernible in our study suggests, on the one hand, that excessive working hours may have a health effect in the German work context. On the other hand, methodological peculiarities could also explain differences from other studies (see below).

Our study also showed a cross-sectional correlation between long commuting times of 10 hours or more and depressive symptoms. However, no effect could be determined in the longitudinal model. Results of international studies on the effect of commuting have been contradictory so far. A German cross-sectional study with more than 4 000 employees of an industrial company found no effects of commuting time on mental health, measured by the SF-12 (20). However, this study was limited to one company, so it is only comparable with our sample to a limited extent. An Australian longitudinal study with more than 17 000 Australian employees found small longitudinal effects of impaired mental health associated with commuting more than 10 hours per week (19). It is possible that these small effects were uncovered due to the statistical power of this much larger study compared to S-MGA. By contrast, a representative longitudinal study of almost 18 000 employees in the UK found no overall effects of longer commuting times on mental health as measured by the PHQ-12 (19). The results of the British study also suggest that the mode of transport used is an important factor: commuting times by foot or by bicycle had a positive effect on mental health, commuting times by car had a negative effect and commuting times by public transport had no effect. A recent review that looked at the role of the mode of transport also shows divergent results (41). Furthermore, avoidance effects are conceivable, such that many employees who feel acutely burdened by commuting times reduce their commuting or switch to other means of transport, so that correlations can no longer be shown longitudinally.

### Methodical Considerations

This study has several strengths, for example, that it is based on a representative sample of employees subject to social insurance contributions (24) and, thus, allows conclusions to be drawn about a large proportion of employees in Germany. Furthermore, the cohort design allows longitudinal analyses and, thus, statements on potential cause-effect relationships or the temporal sequence of exposure and outcome (40, 42). Another advantage is the measurement of actual hours worked, which reflects the exposure better than information on hours worked according to the employment contract. In addition, depressive symptoms were recorded with an established instrument (PHQ-9), the threshold value of which indicates a clinically significant increase (29).

There are a number of limitations to this. One point is that all data are based on self-reports and no objective working and commuting times were recorded over a longer period of time. A bias in self-reports in which both exposures and outcomes are recorded together is, thus, possible, although this problem is likely to be minor in the longitudinal analysis with a five-year interval between measurements (42). Another limitation is that the five-year interval between exposure and outcome measurement is comparatively large. Shorter follow-up times of one to two years would be beneficial (22). Depressive symptoms can change over time and the study design used cannot detect those that appeared and disappeared during the five-year study period (22). In addition, the study design with currently only one baseline survey and one follow-up survey does not permit a closer examination of possible changes over time, for example, whether working or commuting times were reduced in the course of the follow-up due to symptoms at the baseline survey (40). For these reasons, both longitudinal and cross-sectional analyses were conducted in the present study in order to be able to examine at least short-term effects (23). An additional limitation of the present study is – as has already been mentioned above – that the means of transport used for commuting were not recorded. Finally, it should be noted that the self-employed, who often have excessively long working hours, were missing from the sample.

### Perspective

Excessive working hours and commuting times are possible risk factors for depressive symptoms among employees in Germany. Although, according to the current legal situation, excessive working hours should not occur or only in exceptional cases, de facto, a part of the workforce is still affected. In order to avoid adverse health effects, the implementation of existing rules should continue to be consistently promoted. This also applies to the increased option of working from home in the course of the COVID-19 pandemic, which could tempt people to extend their working hours further. Moreover, risk factors can also be taken into account in everyday clinical life, for example, in the context of an occupational history in patients with early symptoms or in those with chronic mental illnesses who continue to work. However, further empirical work on the topic is necessary due to the methodological limitations of this study.

## Data Availability

Access to the S-MGA data is possible through a scientific use file after an application procedure via the Research Data Centre of the Federal Institute for Occupational Safety and Health (doi: 10.21934/baua:doku20210208 [data documentation]; doi: 10.48697/smga.w1w2.suf.1 [Dataset]).

## Acknowledgements

The analyses of the present study were carried out based on the data of the first and second wave of the S-MGA. This panel study was initiated by the Federal Institute for Occupational Safety and Health and conducted in co-operation with the Institute for Employment Research (IAB) and the Institute for Applied Social Science (infas). The sample basis for the S-MGA is the Employment History (BeH) as part of the Integrated Employment Biographies (IEB) of the Federal Employment Agency (BA). Access to the S-MGA data is possible through a scientific use file after an application procedure via the Research Data Centre of the Federal Institute for Occupational Safety and Health (doi: 10.21934/baua:doku20210208 [data documentation]; doi: 10.48697/smga.w1w2.suf.1 [Dataset]).

## Supplementary Tables. Online Appendix

**Supplementary Table A1.** Associations between working and commuting times and depressive symptoms in 3 413 employees in the cross-sectional analyses of the S-MGA, separated by gender (logistic regression; odds ratios (OR) and 95 % confidence intervals (CI)); sensitivity analysis.

|  | Men (n = 1 697) |  |  |  |  | Women (n = 1 716) |  |  |  |  |
| --- | --- | --- | --- | --- | --- | --- | --- | --- | --- | --- |
|  | N | Prevalence of depressive symptoms 2012 (in %) | p | OR* | 95 % CI* | N | Prevalence of depressive symptoms 2012 (in %) | p | OR* | 95 % CI* |
| Weekly working time (hours) |  |  | 0.001 |  |  |  |  | .030 |  |  |
| 10 to < 35 | 51 | 16 |  | 5.76 | 2.54;13.07 | 785 | 7 |  | 0.65 | 0.43;0.98 |
| 35 to < 41 | 609 | 5 |  | 1 |  | 504 | 11 |  | 1 |  |
| 41 to < 49 | 622 | 6 |  | 1.54 | 0.91;2.58 | 303 | 14 |  | 1.20 | 0.75;1.92 |
| 49 to < 55 | 216 | 6 |  | 1.91 | 0.93;3.92 | 73 | 11 |  | 1.60 | 0.74;3.45 |
| ≥ 55 | 199 | 2 |  | 1.80 | 0.87;3.70 | 51 | 21 |  | 1.47 | 0.59;3.64 |
| Weekly commuting time (hours) |  |  | 0.083 |  |  |  |  | .481 |  |  |
| < 2.5 | 471 | 4 |  | 1 |  | 600 | 10 |  | 1 |  |
| 2.5 to < 5 | 635 | 6 |  | 1.19 | 0.69;2.07 | 633 | 10 |  | 1.15 | 0.78;1.71 |
| 5 to < 10 | 444 | 6 |  | 1.47 | 0.82;2.63 | 382 | 9 |  | 0.92 | 0.57;1.48 |
| ≥ 10 | 147 | 17 |  | 2.40 | 1.19;4.84 | 101 | 12 |  | 1.52 | 0.77;3.00 |

\* Adjusted for age, occupational position, psychosocial working conditions. Working and commuting time not mutually adjusted.

**Supplementary Table A2.** Associations between working and commuting times at baseline and depressive symptoms at follow-up of 2 019 S-MGA employees, separated by gender (logistic regression; odds ratios (OR) and 95 % confidence intervals (CI)); sensitivity analysis.

|  | Men (n = 976) |  |  |  |  | Women (n = 1 043) |  |  |  |  |
| --- | --- | --- | --- | --- | --- | --- | --- | --- | --- | --- |
|  | N | Prevalence of depressive symptoms 2017 (in %) | p | OR* | 95 % CI* | N | Prevalence of depressive symptoms 2017 (in %) | p | OR* | 95 % CI* |
| Weekly working time (hours) |  |  | 0.385 |  |  |  |  | .115 |  |  |
| 10 to < 35 | 25 | 12 |  | 1.81 | 0.46;7.07 | 471 | 10 |  | 0.80 | 0.34;1.94 |
| 35 to < 41 | 364 | 6 |  | 1 |  | 309 | 13 |  | 1 |  |
| 41 to < 49 | 356 | 8 |  | 1.62 | 0.87;3.01 | 182 | 15 |  | 1.21 | 0.92;1.83 |
| 49 to < 55 | 125 | 5 |  | 1.06 | 0.39;2.89 | 47 | 19 |  | 1.81 | 0.68;2.13 |
| ≥ 55 | 106 | 8 |  | 2.13 | 0.85;5.34 | 34 | 27 |  | 2.49 | 0.97;4.97 |
| Weekly commuting time (hours) |  |  | 0.882 |  |  |  |  | .193 |  |  |
| < 2.5 | 263 | 6 |  | 1 |  | 350 | 15 |  | 1 |  |
| 2.5 to < 5 | 365 | 7 |  | 1.03 | 0.59;1.94 | 393 | 12 |  | 0.74 | 0.46;1.17 |
| 5 to < 10 | 258 | 7 |  | 1.00 | 0.72;1.64 | 240 | 2 |  | 0.76 | 0.44;1.30 |
| ≥ 10 | 90 | 9 |  | 1.41 | 0.65;3.01 | 60 | 7 |  | 0.32 | 0.11;1.00 |

\* Adjusted for depressive symptoms at baseline, age, occupational position, psychosocial working conditions. Working and commuting time not mutually adjusted.

**Supplementary Table A3.**
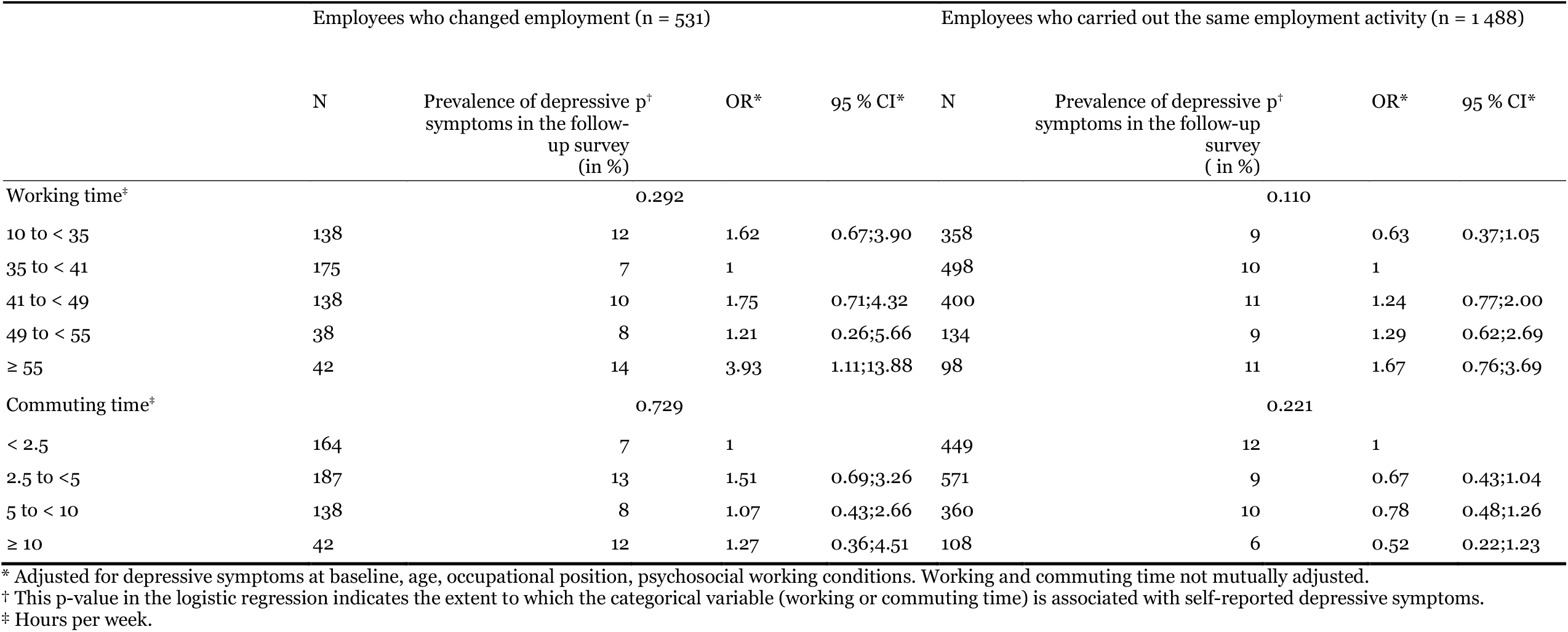
Separated according to the whereabouts or change of employment during follow-up: associations between working and commuting times in the baseline survey and depressive symptoms in the follow-up survey of 2 019 employees of the S-MGA (logistic regression; odds ratios (OR) and 95 % confidence intervals (CI)); sensitivity analysis.

**Supplementary Table A4.**
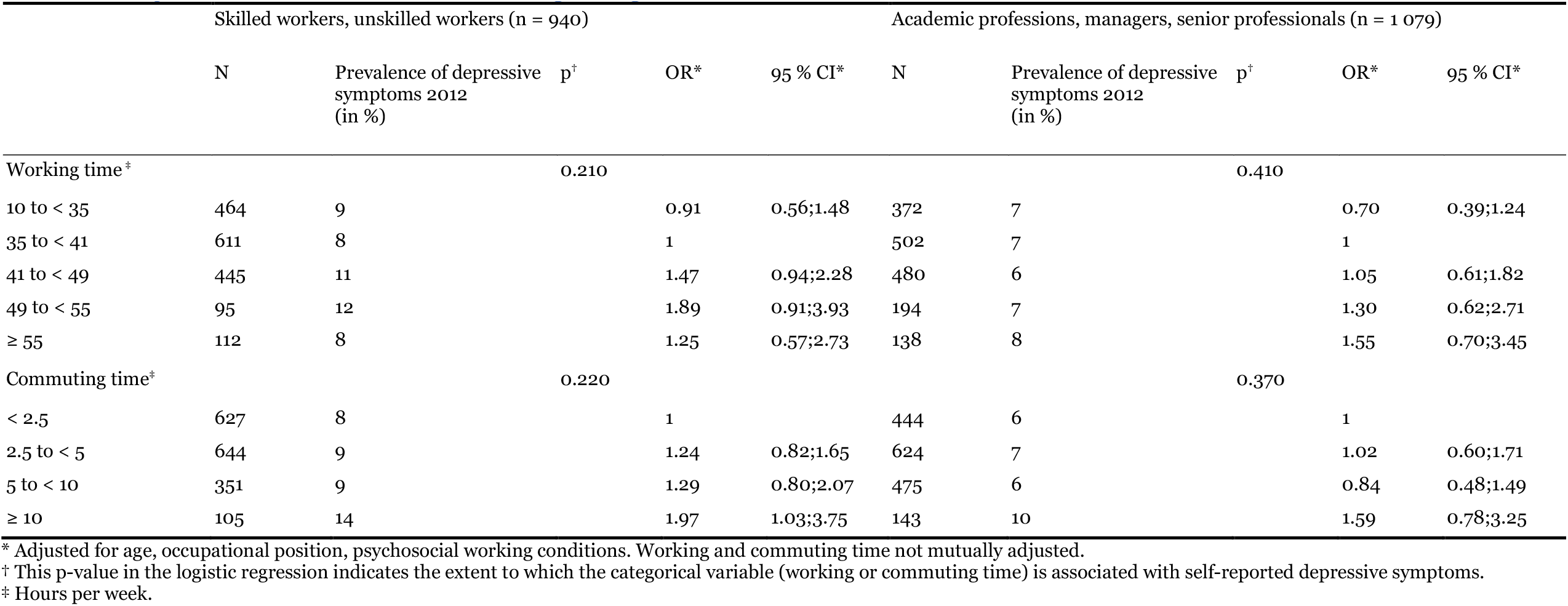
Separated by occupational status: associations between working and commuting times and depressive symptoms in 3 413 employees in the cross-sectional analyses of the S-MGA (logistic regression; odds ratios (OR) and 95 % confidence intervals (CI)); sensitivity analysis.

**Supplementary Table A5.**
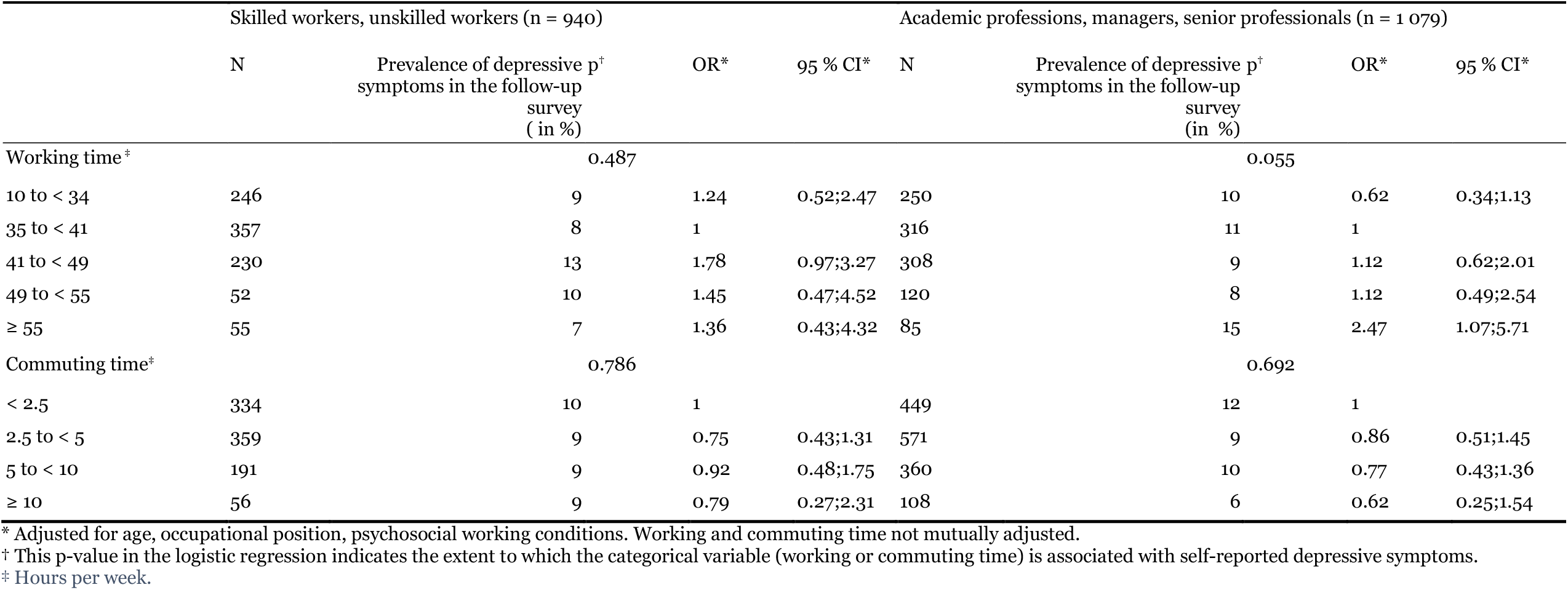
Separated by occupational status: associations between working and commuting times in the baseline survey and depressive symptoms in the follow-up survey of 2 019 employees of the S-MGA (logistic regression; odds ratios (OR) and 95 % confidence intervals (CI)); sensitivity analysis.

## References

1. Deutsche Rentenversicherung Bund: Rentenversicherung in Zeitreihen. Berlin: DRV; 2019.

2. Angerer P, Glaser J, Gündel H, et al.: Psychische und psychosomatische Gesundheit in der Arbeit: Wissenschaft, Erfahrungen und Lösungen aus Arbeitsmedizin, Arbeitspsychologie und Psychosomatischer Medizin. Heidelberg, München, Landsberg, Frechen, Hamburg: ecomed-Storck GmbH; 2014.

3. Bundesministerium für Arbeit und Soziales (BMAS): Arbeitszeitgesetz. In: Verbraucherschutz BfJuf, (ed.). Berlin 2020.

4. Virtanen M, Jokela M, Madsen IE, et al.: Long working hours and depressive symptoms: systematic review and meta-analysis of published studies and unpublished individual participant data. Scandinavian journal of work, environment & health 2018; 44: 239–50.

5. Rugulies R, Sørensen K, Di Tecco C, et al.: The effect of exposure to long working hours on depression: A systematic review and meta-analysis from the WHO/ILO Joint Estimates of the Work-related Burden of Disease and Injury. Environment International 2021; 155: 106629.

6. Backhaus N, Tisch A, Wöhrmann AM: BAuA-Arbeitszeitbefragung: Vergleich 2015 – 2017 – 2019. Dortmund: Bundesanstalt für Arbeitsschutz und Arbeitsmedizin; 2020.

7. Statistisches Bundesamt: Überlange Arbeitszeiten https://www.destatis.de/DE/Themen/Arbeit/Arbeitsmarkt/Qualitaet-Arbeit/Dimension-3/ueberlange-arbeitszeiten.html2020).

8. Bundesministerium für Arbeit und Soziales (BMAS): Verordnung über die Arbeitszeit der Beamtinnen und Beamten des Bundes In: Verbraucherschutz BdJuf, (ed.): § 87 Abs 3 S 1, § 90 Abs 1 BBG. Berlin: Bundesministeriums der Justiz und für Verbraucherschutz 2020.

9. Statistisches Bundesamt [destatis]: Ein Drittel der Ärztinnen und Ärzte arbeitete 2018 mehr als 48 Stunden pro Woche. Pressemitteilung Nr N 019. Berlin: Statistisches Bundesamt [destatis] 2020.

10. Sonnentag S: The recovery paradox: Portraying the complex interplay between job stressors, lack of recovery, and poor well-being. Research in Organizational Behavior 2018; 38: 169–85.

11. Virtanen M, Jokela M, Nyberg ST, et al.: Long working hours and alcohol use: systematic review and meta-analysis of published studies and unpublished individual participant data. Bmj 2015; 350: g7772.

12. Backhaus N, Brauner C, Tisch A: Auswirkungen verkürzter Ruhezeiten auf Gesundheit und Work-Life-Balance bei Vollzeitbeschäftigten: Ergebnisse der BAuA-Arbeitszeitbefragung 2017. Zeitschrift für Arbeitswissenschaft 2019; 73: 394–417.

13. Müller G, Tisch A, Wöhrmann AM: The impact of long working hours on the health of German employees. German Journal of Human Resource Management 2018; 32: 217–35.

14. Wöhrmann AM, Brenscheidt F, Gerstenberg S: Arbeitszeit in Deutschland: Länge, Lage, Flexibilität der Arbeitszeit und die Gesundheit der Beschäftigten. In: Rump J, Eilers S, (eds.): Arbeitszeitpolitik Zielkonflikte in der betrieblichen Arbeitszeitgestaltung lösen. Berlin, Heidelberg: Springer Gabler 2019; p. 159–77.

15. Garthus-Niegel S, Hegewald J, Seidler A, et al.: The Gutenberg health study: associations between occupational and private stress factors and work-privacy conflict. BMC public health 2016; 16: 192.

16. Pega F, Momen NC, Ujita Y, Driscoll T, Whaley P: Systematic reviews and meta-analyses for the WHO/ILO Joint Estimates of the Work-related Burden of Disease and Injury. Environment International 2021; 155: 106605.

17. Dragano N, Siegrist J, Wahrendorf M: Welfare regimes, labour policies and unhealthy psychosocial working conditions: a comparative study with 9917 older employees from 12 European countries. Journal of epidemiology and community health 2011; 65: 793–9.

18. Wöhrmann AM, Backhaus N, Tisch A, Michel A: BAuA-Arbeitszeitbefragung: Pendeln, Telearbeit, Dienstreisen, wechselnde und mobile Arbeitsorte. Bundesanstalt für Arbeitsschutz und Arbeitsmedizin BAuA, Dortmund 2020.

19. Milner A, Badland H, Kavanagh A, LaMontagne AD: Time Spent Commuting to Work and Mental Health: Evidence From 13 Waves of an Australian Cohort Study. The American Journal of Epidemiology 2017; 186: 659–67.

20. Mauss D, Jarczok MN, Fischer JE: Daily commuting to work is not associated with variables of health. Journal of occupational medicine and toxicology 2016; 11: 12.

21. Martin A, Goryakin Y, Suhrcke M: Does active commuting improve psychological wellbeing? Longitudinal evidence from eighteen waves of the British Household Panel Survey. Preventive medicine 2014; 69: 296–303.

22. Ford MT, Matthews RA, Wooldridge JD, Mishra V, Kakar UM, Strahan SR: How do occupational stressor-strain effects vary with time? A review and meta-analysis of the relevance of time lags in longitudinal studies. Work & Stress 2014; 28: 9–30.

23. Boini S, Kolopp M, Grzebyk M, Hédelin G, Chouanière D: Is the effect of work-related psychosocial exposure on depressive and anxiety disorders short-term, lagged or cumulative? International archives of occupational and environmental health 2020; 93: 87–104.

24. Rose U, Schiel S, Schroder H, et al.: The Study on Mental Health at Work: Design and sampling. Scandinavian journal of public health 2017; 45: 584–94.

25. American Association for Public Opinion Research: Standard definitions: Final dispositions of case codes and outcome rates for surveys. 7th ed 2011.

26. Feveile H, Olsen O, Hogh A: A randomized trial of mailed questionnaires versus telephone interviews: response patterns in a survey. BMC medical research methodology 2007; 7: 27.

27. Kroenke K, Spitzer RL, Williams JB: The PHQ-9: validity of a brief depression severity measure. Journal of General Internal Medicine 2001; 16: 606–13.

28. Löwe B, Spitzer RL, Gräfe K, et al.: Comparative validity of three screening questionnaires for DSM-IV depressive disorders and physicians’ diagnoses. Journal of affective disorders 2004; 78: 131–40.

29. Manea L, Gilbody S, McMillan D: A diagnostic meta-analysis of the Patient Health Questionnaire-9 (PHQ-9) algorithm scoring method as a screen for depression. General Hospital Psychiatry 2015; 37: 67–75.

30. Kivimaki M, Jokela M, Nyberg ST, et al.: Long working hours and risk of coronary heart disease and stroke: a systematic review and meta-analysis of published and unpublished data for 603,838 individuals. Lancet (London, England) 2015; 386: 1739–46.

31. Busch MA, Maske UE, Ryl L, Schlack R, Hapke U: [Prevalence of depressive symptoms and diagnosed depression among adults in Germany: results of the German Health Interview and Examination Survey for Adults (DEGS1)]. Bundesgesundheitsblatt, Gesundheitsforschung, Gesundheitsschutz 2013; 56: 733–9.

32. Schmitt M, Altstötter-Gleich C, Hinz A, Maes J, Brähler E: Normwerte für das Vereinfachte Beck-Depressions-Inventar (BDI-V) in der Allgemeinbevölkerung [Norm values for the simplified Beck Depression Inventory (BDI-V) in the general population]. Diagnostica 2006; 52: 8.

33. Hagen F: Levels of Education: Relation between ISCO Skill Level and ISCED Categories http://www.fernunihagen.de/FTB/telemate/database/isced.htm#ISCO (last accessed on Mach 21, 2015.

34. International Labor Office Staff: International Standard Classification of Occupations 2008 (ISCO-08): Structure, Group Definitions and Correspondence Tables. Geneva, Switzerland: International Labour Office; 2012.

35. Müller W, Wirth H, Bauer G, Pollak R, Weiss F: Entwicklung einer europäischen sozioökonomischen Klassifikation. Wirtschaft und Statistik 2007: 527–30.

36. Nübling M, Stößel U, Hasselhorn H-M, Michaelis M, Hofmann F: Measuring psychological stress and strain at work - Evaluation of the COPSOQ Questionnaire in Germany. Psycho-social medicine 2006; 3: Doc05.

37. Karasek R, Theorell T: Healthy work : stress, productivity, and the reconstruction of working life. New York, NY, USA: Basic Books; 1990.

38. Andersson T, Alfredsson L, Kallberg H, Zdravkovic S, Ahlbom A: Calculating measures of biological interaction. European Journal of Epidemiology 2005; 20: 575–9.

39. Burr H, Lange S, Freyer M, et al.: Physical and psychosocial working conditions as predictors of 5-year changes in work ability among 2078 employees in Germany. International archives of occupational and environmental health 2022; 95: 153–68.

40. Taris TW, Kompier MAJ: Cause and effect: Optimizing the designs of longitudinal studies in occupational health psychology. Work & Stress 2014; 28: 1–8.

41. Marques A, Peralta M, Henriques-Neto D, Frasquilho D, Rubio Gouveira É, Gomez-Baya D: Active Commuting and Depression Symptoms in Adults: A Systematic Review. International journal of environmental research and public health 2020; 17.

42. Zapf D, Dormann C, Frese M: Longitudinal studies in organizational stress research: a review of the literature with reference to methodological issues. Journal of Occupational Health Psychology 1996; 1: 145–69.

43. Andersson T, Alfredsson L, Källberg H, Zdravkovic S, Ahlbom A: Excel sheet to calculate measures of biological interaction https://www.researchgate.net/publication/342077886_August_26_2020_Alternative_working_link_to_referred_excel_file_httpswwwbiostatistikseepinetcalculationxls (last accessed on December 9, 2020 2020).

44. Rothman K: Epidemiology. An introduction. New York, NY, USA: Oxford University Press; 2002.

